# The impact of COVID-19 restrictions on older adults’ loneliness: Evidence from high-frequency panel data in Austria

**DOI:** 10.1101/2021.12.15.21267860

**Authors:** Erwin Stolz, Hannes Mayerl, Wolfgang Freidl

## Abstract

**BACKGROUND:** It is unclear how strong and long lasting the effects of (recurring) COVID-19 restrictions are on older adults’ loneliness.

**METHODS:** 469 retired older Austrians (60+) provided 9,732 repeated observations of loneliness across 30 waves of the Austrian Corona Panel Project between March 2020 and March 2022. Ordinal mixed regression models were used to estimate the effect of the strictness of COVID-19 restrictions (stringency index, range=0-100) on older adult’s loneliness.

**RESULTS:** The proportion of older adults who reported to be often lonely correlated (r=0.45) with the stringency index over time: both peaked during lock-downs (stringency index=82, often lonely=10-13%) and were lowest during the summer of 2020 (stringency index=36, often lonely=4-6%). Results from regression models indicate, that when the stringency index increased, loneliness also increased. In turn, as COVID-19 restrictions loosened, levels of loneliness decreased again. Older adults who lived alone were more affected in terms of loneliness by COVID-19 restriction measures compared to those living with others.

**CONCLUSIONS:** More stringent COVID-19 restrictions were associated with an increased in (situational) loneliness among older adults in Austria, and this effect was stronger among those who lived alone. Efforts should be made to enable older adults, in particular those who live alone, to have save in-person contact during periods of strict pandemic restriction measures.

## Introduction

After two years, the COVID-19 pandemic still represents an acute global health threat, with older adults particularly at risk[1]. To break waves of exponential COVID-19 infection rates and to avoid overburdening hospital care, many European governments repeatedly responded with an array of statutory containment and closure policies[2]. By restricting in-person social contacts, however, these public health interventions may lead to negative psychosocial side effects among older adults such as loneliness, which can be defined as a perceived discrepancy between the desired and one’s existing social relationships[3]. Already before the pandemic, older adults had an elevated risk for loneliness due to loss of partners and peers or health-problems[4–7], and loneliness has long been considered harmful to older adults’ physical and mental health[8–10]. Therefore, loneliness induced by pandemic restrictions is currently a subject of concern.

Whether and how strongly COVID-19 restrictions have affected older adults is not yet clear though. While some studies[11–16] suggest an increase in loneliness during the first wave of the pandemic compared to pre-pandemic times, others[17–20] found no changes. We know even less about how loneliness levels changed after the first lock-down. Several studies with multiple measurement points during the early pandemic[13,16,21,22] suggest that loneliness was higher during and immediately after the first lock-down, but leveled off thereafter. In contrast, a small longitudinal study from the US and Canada[23] found stable loneliness during the first five months of the pandemic, and a study from Norway[20] found that while there were no changes between the time before the pandemic and it’s first wave, loneliness increased strongly among older adults during the second wave in late 2020. Importantly, the stringency of pandemic restrictions is not explicitly measured in these studies – although it’s variation between countries and over time has been considerable – and has not been differentiated from the pandemic itself, i.e. the incidence rate of COVID-19 infections, which may cause voluntary social distancing above and beyond instituted public health restriction measures.

Living alone is a known risk factor for loneliness[5,7], and older adults who live alone could be particularly affected by COVID-19 related restrictions that limit in-person contacts with individuals from outside the household. In comparison, older adults who live through phases of restricted social activities or strict lock-downs together with their co-residing partner or children should be less affected with regard to loneliness. Indeed, a few studies reported that during the early pandemic, increases in loneliness were higher among older adults who lived alone[11,16,24] or were un-partnered[12,20], but it is unclear whether these findings extend throughout the pandemic. In sum, we currently know rather little about later periods of the pandemic: It is unclear how strong and long lasting the effects of COVID-19 restrictions on older adults’ loneliness are, and whether those who live alone were particularly affected. Existing research on this topic is hampered by retrospective survey questions, cross-sectional data, or, in case of longitudinal studies, by long periods between few repeated observations, which’ timing may not align well with waves of COVID-19 infections and instituted lock-downs. Finally, most studies lack quantitative measures of the exposure, i.e. the pandemic-related restrictions, and are therefore unable to establish whether a dose-response relationship with loneliness exists. In the current study, we attempt to improve upon these issues by exploiting high-frequency panel data from Austria, which allows to monitor older adults continuously throughout the entire pandemic in order to assess the real-time impact of the stringency of COVID-19 restrictions on loneliness.

## Methods

### Data

For this study, we used data from the Austrian Corona Panel Project[25], a high-frequency online panel survey of the general population in Austria with >1,500 participants per wave conducted by the University of Vienna. Between March 27th 2020 and March 25th 2022, 30 waves of online interviews – initially on a weekly basis, then approximately once per month – have been completed. To participate, respondents had to be Austrian residents and aged 14 years or older. Respondents were quota sampled from a pre-existing online panel based on key demographics (age, gender, region, municipality size, educational level) closely mirroring the Austrian resident population. The initial participation rate was 35%, and the retention rates for panelists ranged from 86% in wave 2 to 48% in wave 30. For our current study on older adults’ loneliness, we used data from the 469 retired participants aged 60 years and over who provided a total of 9,732 repeated measurements (=23 interviews per person on average).

### Variables

Loneliness was measured with the same single item in each wave: participants were asked how often they felt lonely during the last week. Possible answer categories included “never”, “on some days”, “multiple times a week”, “almost every day”, and “every day”.

Individual-level predictor variables included the time of interview since baseline (in weeks), and five time-invariant variables: living alone (no/yes), age (in years), sex (male/female), high school education (no/yes), and having one or more of the following chronic disease(s): cardiovascular disease, diabetes, hepatitis B, chronic obstructive lung disease, chronic kidney disease or cancer (no/yes). 12 participants (2.6%) had missing values in these variables, which was addressed by multiple imputation (5 imputated data sets).

To measure the stringency of pandemic-related restrictions, we used the COVID-19 Government Response Stringency Index (SI, [26]) as a time-varying, country-level predictor. The stringency index is a sum index based on nine ordinal measures (school closing, workplace closing, canceling of public events, restriction on gathering size, public transport closing, stay at home requirements, restrictions on internal movement, international travel control, and public information campaigns) that quantifies pandemic-related containment and closure policies on a daily basis, ranging from 0 (no restrictions) to 100 (maximum restrictions). Since the loneliness item refers to the last week before each interview, we calculated lagged 7-day smoothed stringency index values. As an additional country-level and time-varying predictor, we included the number of new COVID-19 cases in Austria (lagged 7-day smoothed values, per 1000; Source: OurWorldInData).

### Statistical Analysis

First, we analysed loneliness and the stringency of COVID-19 restrictions descriptively on the aggregate level by plotting the prevalence of categories of loneliness for each survey wave alongside the stringency index across time. We then calculated the Pearson correlation coefficient between the SI and the prevalence of loneliness categories across waves (n=30). For easier presentation, we collapsed the last three categories of loneliness (multiple days a week, almost every day, and every day) here to “often lonely”.

Second, we modeled the impact of the time-varying SI on repeatedly measured individual-level loneliness using generalized mixed regression models. Specifically, we fitted cumulative ordinal logistic regression models[27] to handle the five non-equidistant response categories and the strong positive skew of the outcome[28]. In the first model, we included time in weeks as a linear predictor to see whether loneliness among older adults has increased during the pandemic. We also included random intercept and slope (week) terms to account for repeated observations nested within respondents. Our core interest, however, was in the overall effect of the time-varying stringency of COVID-19-related restrictions on loneliness. To differentiate the specific effect of restriction measures from the course of the pandemic and its current threat level – which may cause older adults to restrict social contacts above and beyond public restriction measures – we adjusted for the number of new COVID-19 cases. Next to the overall effect of the SI on loneliness, we were also interested whether this is moderated by living alone. Therefore, we added an interaction effect between SI and living alone in the second model. However, since the relationship between living alone and loneliness might be confounded by socio-demographics and health[29,30], we also adjusted for these in model 2. For all analyses, we applied demographic weights.

All analyses were conducted in *R* (v4.1.3): Multiple imputation was performed via chained equations with R-package *mice*. Bayesian mixed ordinal regression models using a Hamiltonian Monte Carlo procedure were estimated with R-package *brms* (v2.16.3)[31], a front-end for *RStan* (v2.21.3). Plots were created with R-package *ggplot2* (v.3.3.5).

## Results

The median age of the sample was 69 (IQR=8, range=60-85) years, 57.8% were women, 16.4% had completed high school education, 42.2% had one or more chronic disease(s), and 32.8% lived alone before the onset of the pandemic. The stringency of COVID-19 restrictions varied considerably during the 2-year pandemic period in Austria (Figure 1): they peaked (SI=82) during the first three lock-down periods (March-April 2020, November-December 2020, January 2021) and were lowest during the summer months of 2020 (SI=36) and 2021 (SI=49). New COVID-19 infections remained low during the first lock-down (<800 cases in March 2020), increased before the second lock-down (<8,000 cases in November 2020), and peaked toward the end of the observation period (>45,000 cases in March 2022).

**Figure 1:**
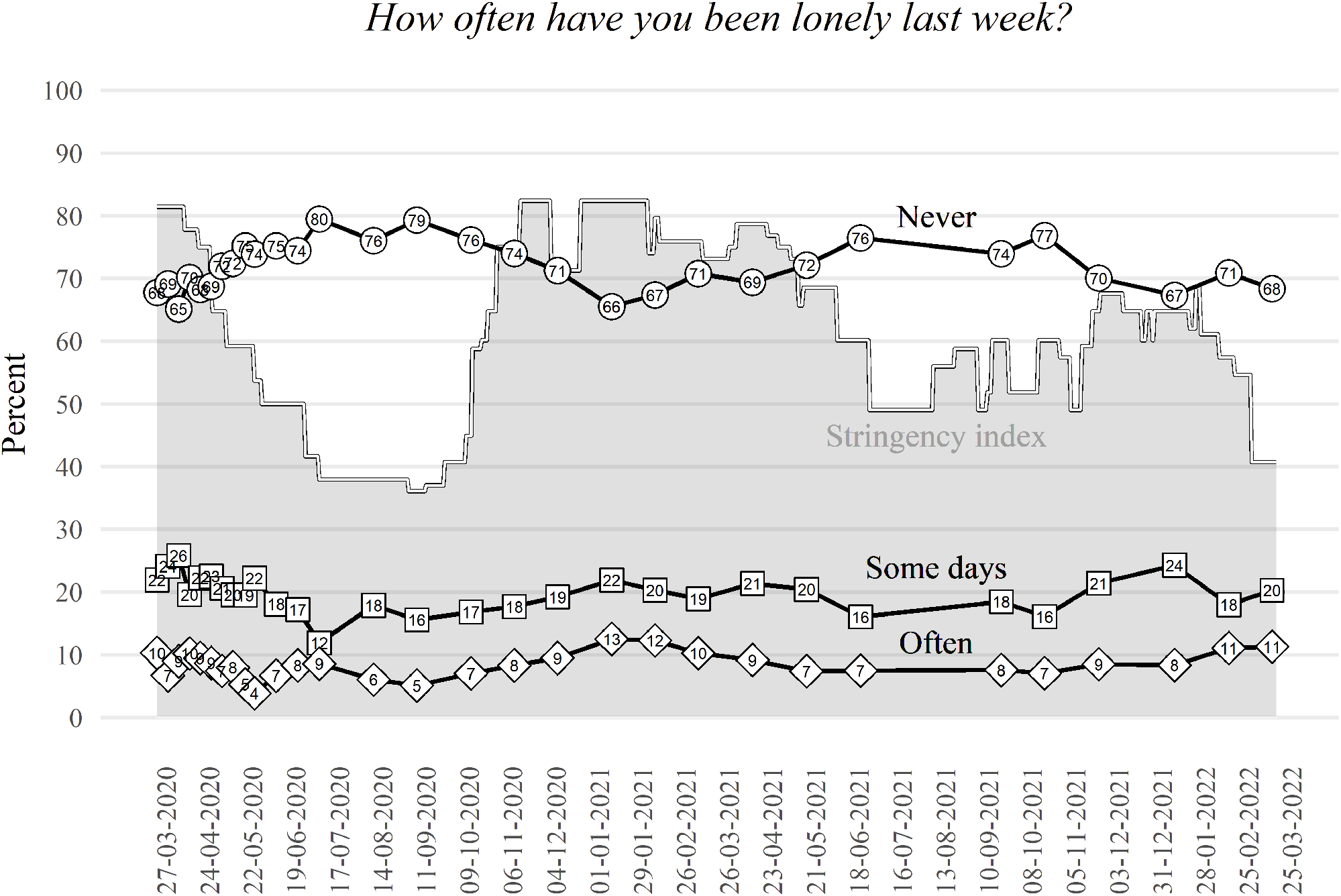
Prevalence of loneliness (March 2020 March 2022) n=469, weighted data. Figure shows trajectories of the prevalence of the categories of loneliness (black lines) in % by wave. Answer categories ‘multiple times a week’, ‘almost every day’, and ‘every day’ were summarised as ‘often lonely’ for easier presentation and interpretation. Grey background shows the stringency index (SI).

Overall, most participants reported to be ‘never’ lonely (72.6%), or to be lonely only on ‘some days’ (19.6%). Few reported to be lonely more often: 3.8% reported to be lonely ‘multiple days a week’, 2.2% ‘almost every day’, and 1.9% ‘every day’. However, the prevalence of feeling often lonely (multiple days per week or more often) varied considerably during the course of the pandemic (Figure 1). The proportion of older adults who felt often lonely reached its maximum of 10-13% during lock-down periods, but decreased again thereafter. During the summer months of 2020, when restrictions were lowest, only 4-6% reported to be often lonely. The correlation coefficient between the SI and the prevalence of feeling often lonely was r=0.45 across waves. This pattern, however, varied by living status (Figure 2): Older adults who lived alone were not only considerably more likely to report feeling often lonely on average – maximal proportion was 19-23% during lock-downs – but their loneliness was also more closely tethered to the SI (r=0.48) compared to those who lived together with others (r=0.18).

**Figure 2:**
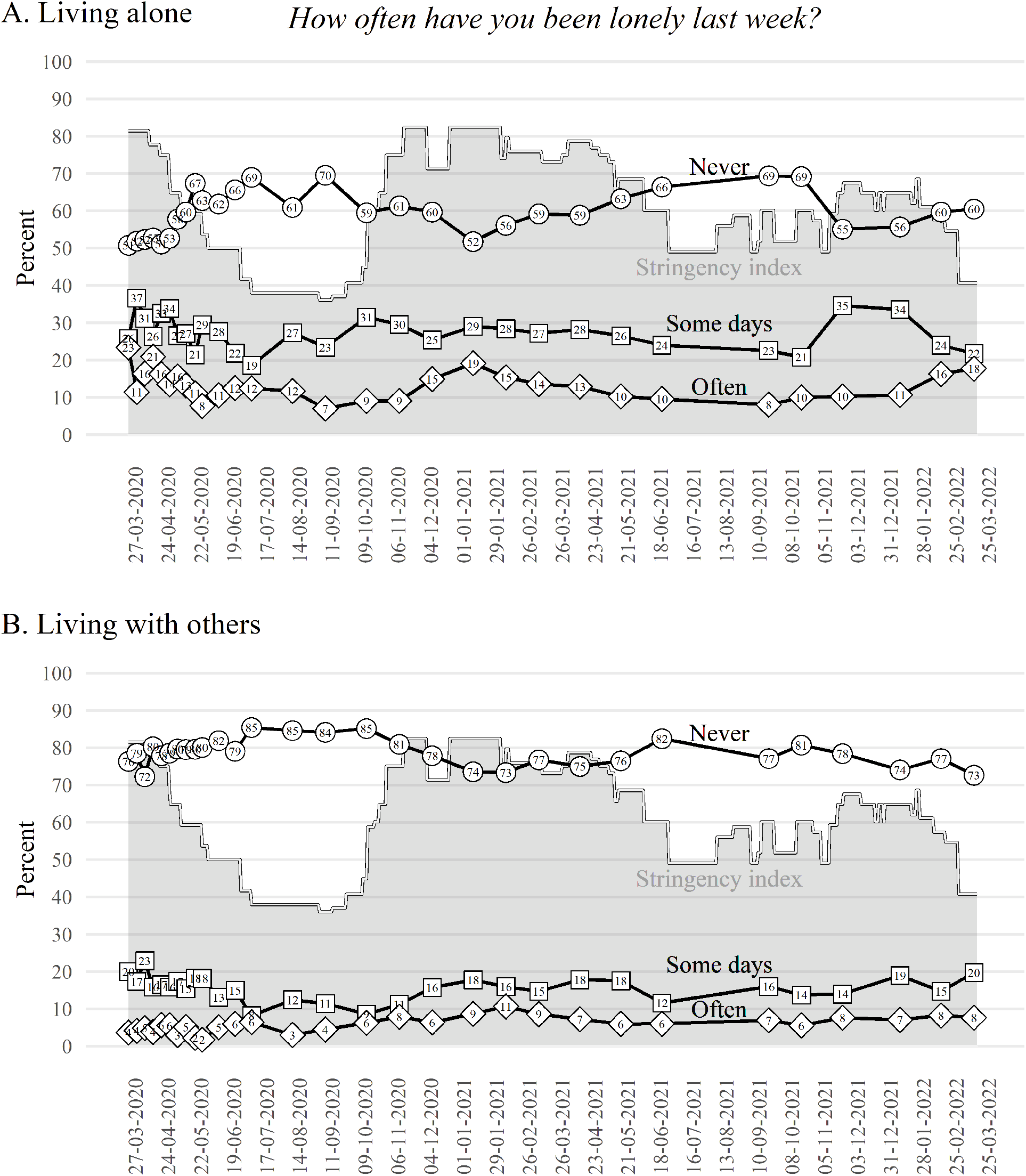
Prevalence of loneliness by living status (March 2020 February 2022) Weighted data. Figure shows trajectories of the prevalence in % of categories of loneliness (black lines) for those who lived alone (plot A, n = 166) and for those who lived with others (plot B, n = 303). Answer categories ‘multiple times a week’, ‘almost every day’, and ‘every day’ were summarised as ‘often lonely’ for easier presentation and interpretation. Grey background shows the SI.

Results from the regression models (Table 1) show no (linear) overall increase in loneliness across the two-year period, but even a slight decrease. Looking at the effect of the SI, we see that a one-point increase (total range in Austria between 2020-2022 was 46 points) was associated with a 3%-increase in the odds of being more lonely (model 1). Specifically, the estimated probability to feel lonely ‘on some days’ increased from 5% when the stringency of COVID-19 restrictions was lowest to 13% when the restrictions were strictest. For the probability of being often lonely, the maximum difference in the restrictions’ stringency during the observed period translates to an associated maximum change from 0.2% to 0.7%.

**Table 1:**
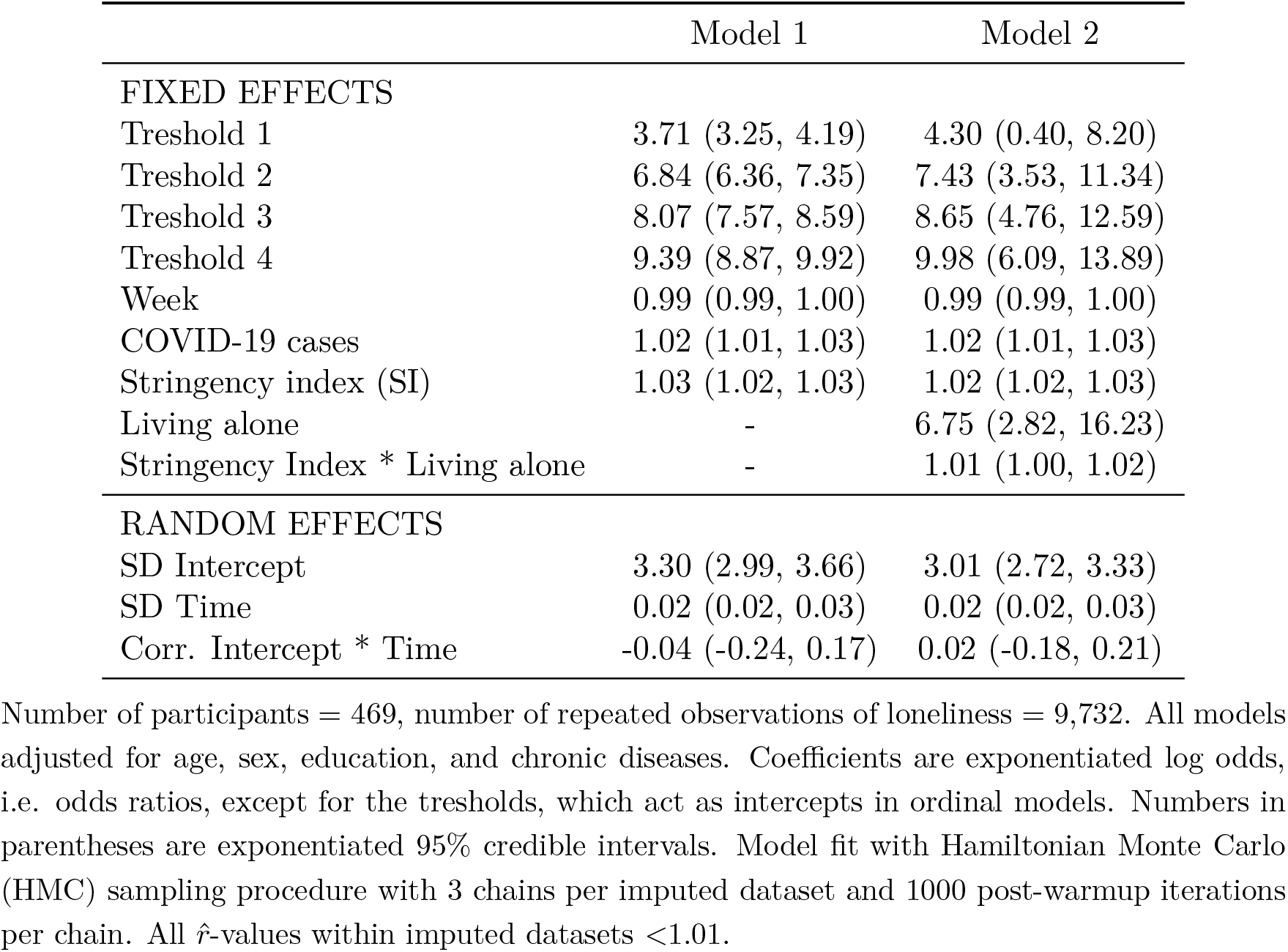
Results from ordinal mixed regression models

|  | Model 1 | Model 2 |
| --- | --- | --- |
| FIXED EFFECTS |  |  |
| Threshold 1 | 3.71 (3.25, 4.19) | 4.30 (0.40, 8.20) |
| Threshold 2 | 6.84 (6.36, 7.35) | 7.43 (3.53, 11.34) |
| Threshold 3 | 8.07 (7.57, 8.59) | 8.65 (4.76, 12.59) |
| Threshold 4 | 9.39 (8.87, 9.92) | 9.98 (6.09, 13.89) |
| Week | 0.99 (0.99, 1.00) | 0.99 (0.99, 1.00) |
| COVID-19 cases | 1.02 (1.01, 1.03) | 1.02 (1.01, 1.03) |
| Stringency index (SI) | 1.03 (1.02, 1.03) | 1.02 (1.02, 1.03) |
| Living alone | - | 6.75 (2.82, 16.23) |
| Stringency Index * Living alone | - | 1.01 (1.00, 1.02) |
| RANDOM EFFECTS |  |  |
| SD Intercept | 3.30 (2.99, 3.66) | 3.01 (2.72, 3.33) |
| SD Time | 0.02 (0.02, 0.03) | 0.02 (0.02, 0.03) |
| Corr. Intercept * Time | -0.04 (-0.24, 0.17) | 0.02 (-0.18, 0.21) |

Number of participants = 469, number of repeated observations of loneliness = 9,732. All models adjusted for age, sex, education, and chronic diseases. Coefficients are exponentiated log odds, i.e. odds ratios, except for the thresholds, which act as intercepts in ordinal models. Numbers in parentheses are exponentiated 95% credible intervals. Model fit with Hamiltonian Monte Carlo (HMC) sampling procedure with 3 chains per imputed dataset and 1000 post-warmup iterations per chain. All $\hat{r}$ -values within imputed datasets $<1.01$ .

From the second model, we can see not only that living alone is a strong risk factor for loneliness during the pandemic, but also that the effect of the SI is moderated by the living arrangement: older adults who lived alone where more likely to feel lonely as pandemic-related restriction measures increased compared to those who lived with others (bottom row of Figure 3). The estimated probability of feeling often lonely tripled from the lowest to the highest level of pandemic restrictions among those who lived alone, while the estimated change was minimal for those older adults who lived together with partner, children or other household members.

**Figure 3:**
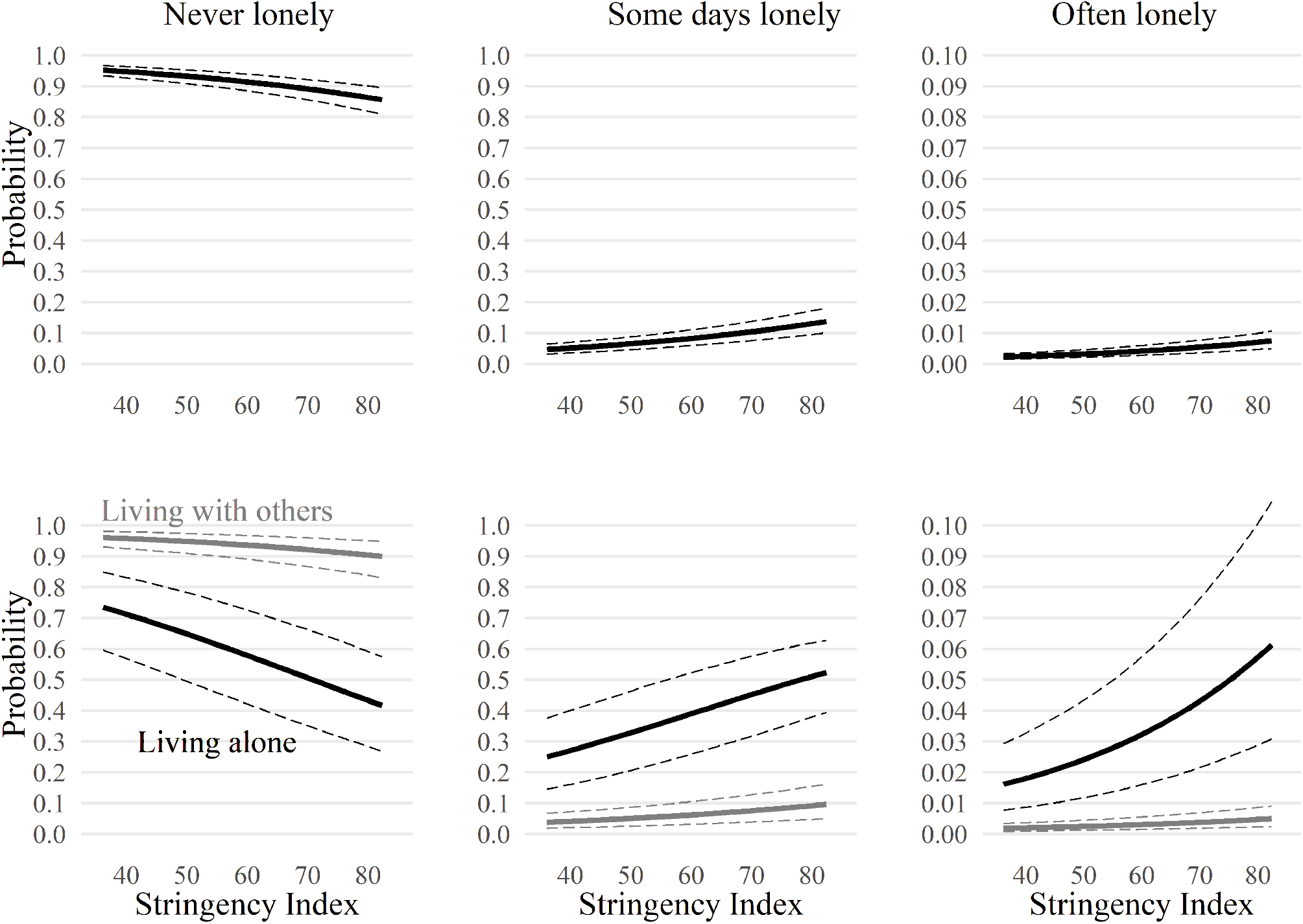
Predicted probability of categories of loneliness by stringency of COVID-19 restrictions Results based on ordinal mixed regression models (first row = model 1, second row = model 2) based on weighted data. Note that the y-axis is not identical between the first two columns and the third column. Estimated probabilities for answer categories ‘multiple times a week’, ‘almost every day’, and ‘every day’ were summed up (often lonely) for easier presentation. Solid lines are predicted probability estimates based on the population-level effects for women of mean age, without high school-level education, and with chronic diseases during the mean point during follow-up. Dashed lines are 95% credible intervals.

## Discussion

In this paper, we monitored loneliness among retired older adults 60+ in Austria throughout two years of the COVID-19 pandemic based on high-frequency panel data in order to assess the real-time impact of COVID-19 restrictions. In summary, we found that when pandemic restrictions became more stringent, the prevalence of loneliness also increased, particularly so among those older adults who lived alone. Soon after restrictions were loosened, however, the prevalence of loneliness tended to decrease again.

The findings of our study are compatible with evidence that loneliness among older adults was higher during the first lock-down in the early pandemic compared to pre-pandemic times[11–16]. Our study extends these findings for later periods of the pandemic in Austria, including multiple subsequent lock-downs and differentiating effects of pandemic-related restrictions from COVID-19 infection rates. Unlike the study from the US and Canada[23], we detected wave-like patterns of loneliness largely congruent with pandemic restrictions, and unlike the study from Norway[20], we found no evidence for strong differences in the effect of restrictions on loneliness between waves or lock-downs. Instead, we confirmed findings of longitudinal studies from the early pandemic[13,16,21,22], insofar that increases in loneliness due to lock-downs seem rather short-lived and reverted after restrictions were lifted. We found no evidence in our study that loneliness generally increased or that it chronified over the course of the COVID-19 pandemic. Therefore, restriction-induced loneliness appears mostly situational. In comparison to chronic loneliness, situational loneliness has been found to be less of a risk factor for negative long-term health outcomes[32,33]. Even so, pandemic-induced loneliness may already have had short-term negative mental health consequences: two studies[15,34], for example, reported pandemic-related loneliness to predict depression and anxiety symptoms among older adults.

The results from our models imply that increases of new COVID-19 infections also had an effect on older adults’ loneliness above and beyond the stringency of pandemic-related cotainment and closure policies introduced by law. This is compatible with evidence from studies[35,36] showing that older adults who voluntarily engaged in physical distancing measures, e.g. avoiding close contact or canceling social activities, also reported higher levels of loneliness.

With regard to the strictness of COVID-19 related restriction measures, we are aware of only one other study[24] that explicitly used the SI as a fine-grained measure of pandemic restrictions and that assessed it as a determinant of older adults’ loneliness. The authors of this pan-European study based on data from retired individuals 60+ in the Survey of Health, Ageing, and Retirement in Europe (SHARE) found no association between the number of days with stringent restriction measures (SI>60) across countries and self-reported changes in loneliness, which contrasts with our findings. This difference to our results, however, could be due to a number of methodological differences: while we modeled the effect of time-varying COVID-19 restrictions on many repeated observations of loneliness in a single country over two years, the pan-European study assessed accumulated differences in stringency across 27 countries at a single, early time point (during summer 2020) based on a retrospective question about pandemic-related change in loneliness. Although it is unclear whether the different results stem from substantive differences – for example, the relationship between the strictness of pandemic-related restrictions and loneliness might vary across countries – or methodological differences (or both), we believe that the analysis of high-frequency longitudinal data in our paper is, in principle, better suited to detect the impact of time-varying COVID-19-related restrictions on older adults’ loneliness given the wave-like pattern of pandemic loneliness we uncovered.

Our results imply an increase in loneliness as restriction measures became more stringent particularly among older adults living alone, who account for 20%-40% of the total older population aged 65+ in Europe and the US[30,37]. These results are in line with findings from the early pandemic, that older adults who lived alone had fewer in-person contacts and provided or received less help from others[38], and that pandemic-related increases in loneliness were higher among them[11,16,24], as well as among those who were un-partnered[12,20]. Therefore, efforts should be made to specifically enable older adults who live alone to have save forms of in-person contact during lock-down periods[38] in order to stay socially connected.

Despite a number of strengths (high-frequency panel data, fine-grained longitudinal measure of COVID-19-related restrictions over time, adjustment for COVID-19 infection rates, and appropriate statistical model for categorical longitudinal data), there are also several limitations to this study. First, we lacked a pre-pandemic baseline rate of loneliness (without any COVID-19 restrictions) for comparison to assess the overall impact of the pandemic. Second, loneliness was measured with a direct, single item: Although frequently used and often highly correlated with established multiple-item scales[7], single items may be less reliable and may also lead to underestimation due to the negative connotations of the term ‘loneliness’. Third, it is unlikely that the quota sample of the current study is truely representative for the population of the general older population in Austria with regard to loneliness – a problem that plagues a large proportion of the current research on this topic[39]. Older adults, particularly the oldest old and institutionalized individuals, and those with a low level of education – all of which are more likely to be lonely[7,29,30], particularly during the current pandemic – are difficult to recruit for online interviews[40]. Despite the use of demographic weights, these limitations likely resulted in an underestimation of the prevalence of loneliness, which may, in consequence, also down-bias our effect estimates on loneliness. In other words, we expect the true effect of COVID-19 restriction measures on older adult’s loneliness to be higher than what we were able to document in the current study.

In conclusion, we found that increases in pandemic restrictions were associated with (situtional) loneliness among older adults in Austria, particularly among those who lived alone.

## Data Availability

Data from the Austrian Corona Panel Project are freely available for scientific research via the Austrian Social Science Data Archive (AUSSDA: https://data.aussda.at/dataset.xhtml?persistentId=doi:10.11587/28KQNS). The R-Markdown code reproducing
all analyses, results and this manuscript is available online via OSF (https://osf.io/aq97s/)

https://osf.io/aq97s/

## Data and code availability

Data from the Austrian Corona Panel Project are freely available for scientific research via the Austrian Social Science Data Archive (AUSSDA). The R-Markdown code reproducing all analyses, results and this manuscript is also available online via OSF.

## Author Contributions

Erwin Stolz is the corresponding author, he planned the study, performed all statistical analysis, and wrote the article. Hannes Mayerl contributed to the interpretation of results and critically reviewed the manuscript. Wolfgang Freidl also critically reviewed the manuscript.

